# Loss-of-function variants in *JPH1* cause congenital myopathy with prominent facial involvement

**DOI:** 10.1101/2024.02.10.24302480

**Authors:** Mridul Johari, Ana Topf, Chiara Folland, Jennifer Duff, Lein Dofash, Pilar Marti, Thomas Robertson, Juan J Vilchez, Anita Cairns, Elizabeth Harris, Chiara Marini-Bettolo, Gianina Ravenscroft, Volker Straub

**Author notes:** Corresponding Authors Gianina Ravenscroft, Harry Perkins Institute of Medical Research, 6 Verdun Street, Nedlands, WA 6009, Australia, Volker Straub, John Walton Muscular Dystrophy Research Centre, Translational and Clinical Research Institute, Newcastle University and Newcastle Hospitals NHS Foundation Trust, Newcastle upon Tyne, UK. These authors contributed equally (Mridul Johari, Ana Topf).

## Abstract

**Background:** Weakness of facial, ocular, and axial muscles is a common clinical presentation in congenital myopathies caused by pathogenic variants in genes encoding triad proteins. Abnormalities in triad structure and function resulting in disturbed excitation-contraction coupling and Ca^2+^ homeostasis can contribute to disease pathology.

**Methods:** We analysed exome and genome sequencing data from three unrelated individuals with congenital myopathy characterised by striking facial, ocular, and bulbar involvement. We collected deep phenotypic data from the affected individuals. We analysed the RNA-seq data of one proband and performed gene expression outlier analysis in 129 samples.

**Results:** The three probands had remarkably similar clinical presentation with prominent facial, ocular, and bulbar features. Disease onset was in the neonatal period with hypotonia, poor feeding, cleft palate and talipes. Muscle weakness was generalised but most prominent in the lower limbs with facial weakness also present. All patients had myopathic facies, bilateral ptosis, ophthalmoplegia and fatiguability. While muscle biopsy on light microscopy did not show any obvious morphological abnormalities, ultrastructural analysis showed slightly reduced triads, and structurally abnormal sarcoplasmic reticulum.

DNA sequencing identified three unique homozygous loss of function variants in *JPH1*, encoding junctophilin-1 in the three families; a stop-gain (c.354C>A; p.Tyr118*) and two frameshift (c.373del p.Asp125Thrfs*30 and c.1738del; p.Leu580Trpfs*16) variants. Muscle RNA-seq showed strong downregulation of *JPH1* in the F3 proband.

**Conclusions:** Junctophilin-1 is critical to the formation of skeletal muscle triad junctions by connecting the sarcoplasmic reticulum and T-tubules. Our findings suggest that loss of JPH1 results in a congenital myopathy with prominent facial, bulbar and ocular involvement.

**Key message:** This study identified novel homozygous loss-of-function variants in the *JPH1* gene, linking them to a unique form of congenital myopathy characterised by severe facial and ocular symptoms. Our research sheds light on the critical impact on junctophilin-1 function in skeletal muscle triad junction formation and the consequences of its disruption resulting in a myopathic phenotype.

**What is already known on this topic:** Previous studies have shown that pathogenic variants in genes encoding triad proteins lead to various myopathic phenotypes, with clinical presentations often involving muscle weakness and myopathic facies. The triad structure is essential for excitation-contraction (EC) coupling and calcium homeostasis and is a key element in muscle physiology.

**What this study adds and how this study might affect research, practice or policy:** This study establishes that homozygous loss-of-function mutations in *JPH1* cause a congenital myopathy predominantly affecting facial and ocular muscles. This study also provides clinical insights that may aid the clinicians in diagnosing similar genetically unresolved cases.

## Introduction

In skeletal muscles, the sarcoplasmic reticulum (SR) is surrounded by specialised invaginations of the sarcolemma in the form of terminal cisternae and transverse tubules (T-tubules). The juxtaposition of a T-tubule with two terminal cisternae forms the triad (1). In the triads, proteins, notably the dihydropyridine receptor (DHPR) in the T-tubule and the ryanodine receptor (RYR) in the SR, maintain Ca^2+^ homeostasis and are crucial in excitation–contraction (EC) coupling (2).

Disturbed EC coupling and Ca^2+^ homeostasis, along with secondary abnormalities of triad structure and function (3) are the pathomechanisms of myopathies associated with variants in genes encoding proteins critical to EC coupling, including *RYR1, CACNA1S, ORAI1, STAC3, STIM1, MTM1, DNM2* and *BIN1*. These disorders are collectively referred to as triadopathies (3).

Junctophilins are key proteins responsible for triad structure formation and maintenance in striated muscle (4).There are three junctophilin genes. *JPH1* is predominantly expressed in skeletal muscles, while *JPH2* is expressed in cardiac and skeletal muscles and *JPH3* specifically in brain (5, 6). In the skeletal muscle triad, JPH1 interacts with RYR1 aiding in the release of Ca^2+^ (Fig. 1a). In vitro, downregulation or loss of Junctophilins can result in defective triads and dysregulated Ca^2+^ homeostasis due to mislocalisation of RYR1 and DHPR (7, 8). *Jph1* knockout mice die shortly after birth, with ultrastructural analysis showing defective and reduced triads along with structurally abnormal SR (4).

**Fig. 1.**
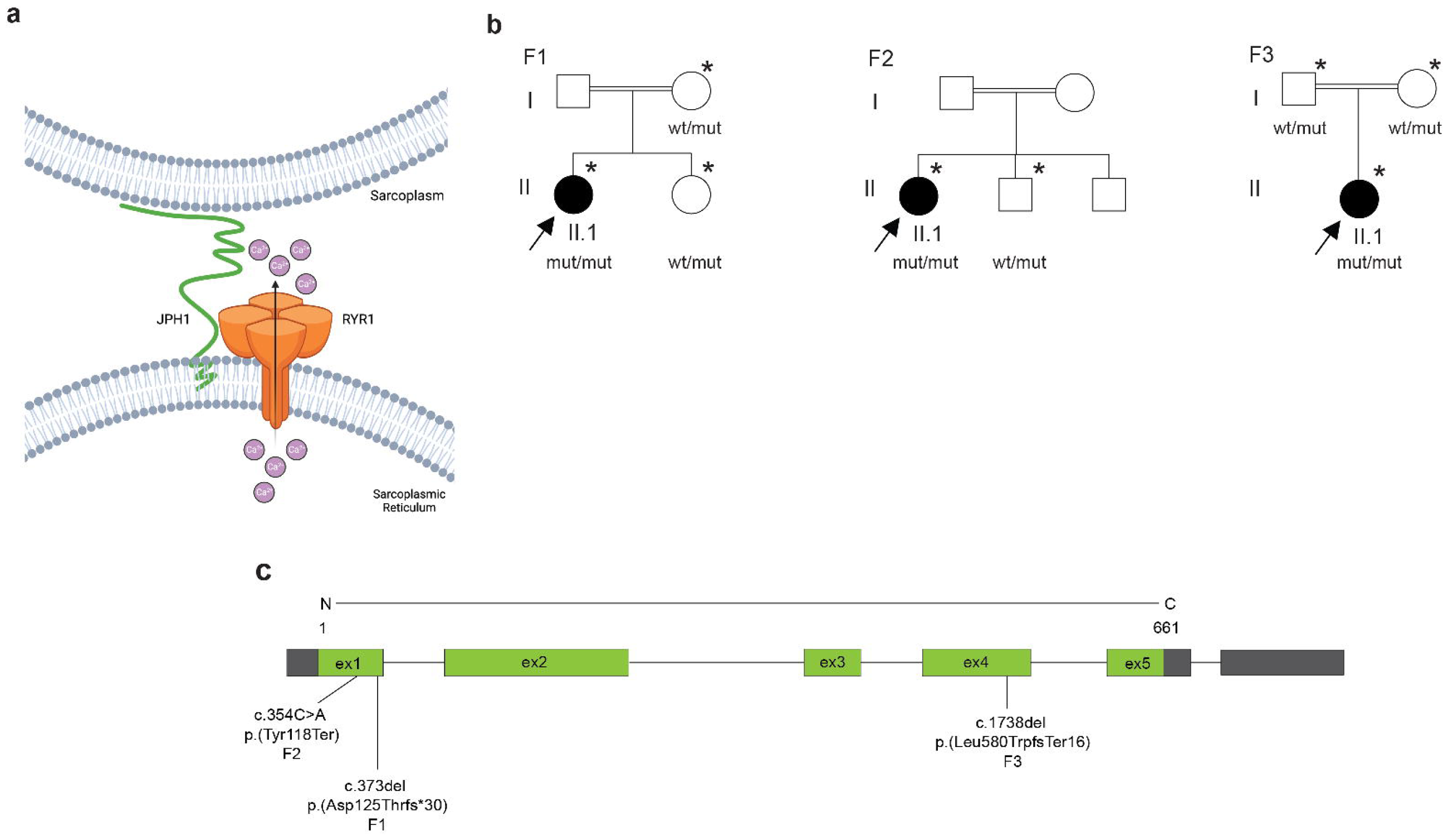
*JPH1*-related myopathy in three families. a) Schematic representation of interaction of JPH1 (Junctophillin-1) and RYR1 (Ryanodine receptor type 1) at the neuromuscular triad. Flow of Ca^2+^ is indicated from the sarcoplasmic reticulum to sarcoplasm through RYR1. b) Pedigrees of the three consanguineous families included in this study, Family 1 and 2 are of European origin, whilst Family 3 is of Asian origin. DNA samples were collected from individuals indicated with ‘*’. The genotype of these individuals is shown as ‘wt/mut’ for heterozygous and ‘mut/mut’ for homozygous for the identified *JPH1* variants. c) A summary of identified pathogenic variants in *JPH1* and their position on the *JPH1* gene model.

Here, we report three unrelated probands with strikingly similar phenotypes involving facial and ocular muscle weakness caused by homozygous null variants in *JPH1*. Our deep phenotyping and novel genetic findings expand the spectrum of congenital myopathies caused by defects in triad proteins and provide evidence for the first time that loss of *JPH1* results in a skeletal muscle disease and should be classified as a triadopathy.

## Methods

### Patients and clinical examinations

Blood samples were collected from three unrelated affected patients and three additional asymptomatic family members. Patient biomaterials for diagnostic purposes were collected after written informed consent was obtained from the patients or their legal guardians by the referring clinicians. The present studies were performed according to the Declaration of Helsinki, and ethical permissions were obtained through institutional review boards including the Human Research Ethics Committee, University of Western Australia, Australia (F3) and the National Research Ethics Service (NRES) Committee North East–Newcastle & North Tyneside 1, United Kingdom (reference 08/H0906/28) (European patients F1 and F2).

All three probands underwent clinical neuromuscular examination. Ancillary tests, including electrophysiological examinations (nerve conduction studies and needle electromyogram, EMG) and serum creatine kinase (CK) levels were obtained in all patients.

### Molecular genetics

Genomic DNA was isolated from blood cells of probands and available family members, using standard techniques.

Exome sequencing (ES) from the genomic DNA of F1-II:1 and F2-II:1 was carried out by the Broad Institute Genomics Platform using an 8-MB targeted Illumina exome capture.

PCR-free libraries were prepared from the genomic DNA of F3-I:1, F3-I:2 and F3-II:1. Shortread (sr) genome sequencing (GS) was performed on NovaSeq 6000 (Illumina, San Diego, CA, USA) with pair-end 150 bp reads at the Kinghorn Centre for Clinical Genomics (Garvan Institute of Medical Research, NSW, Australia).

Single nucleotide variant (SNV) analysis for the three families was performed using *seqr* (9), hosted by the Centre for Population Genomics, a collaboration between Garvan Institute of Medical Research (Sydney, Australia) and the Murdoch Children’s Research Institute (Melbourne, Australia).

ES and srGS results were analysed and SNV/indels were filtered using a minor allele frequency (MAF) ≤ 0.0001 in the Genome Aggregation Database (gnomAD) v2.1.1 (hg19) and v3.1.2 (hg38).

Variants in *JPH1* are annotated on NM_020647.2 and NP_065698.1. All identified variants were also evaluated for current ACMG pathogenicity annotations using VarSome (10), Alamut (Alamut™ Visual Plus version 1.6.1, SOPHiA GENETICS) and Mutalyzer (11).

### Muscle biopsy, immunohistochemical, and imaging studies

Snap-frozen muscle biopsy samples were obtained from the three affected patients. Routine muscle histopathological studies were performed, including hematoxylin & eosin (H&E), modified Gomori’s trichrome, and NADH tetrazolium reductase (NADH-TR) staining (12). DAB immunostaining was performed using mouse monoclonal anti-myotilin (clone RSO34, 1:20, LEICA Biosystems Newcastle Ltd, UK) and mouse monoclonal anti-desmin (clone D33, 1:70, Richard-Allan Scientific, USA), with Mouse ExtrAvidin Peroxidase Staining Kit (EXTRA2, Merck KGaA, Darmstadt, Germany). Microscopic images were obtained using a NIKON ECLIPSE Ci microscope equipped with an OLYMPUS ColorView II camera.

For patient F3-II:1, ultrathin resin sections with a thickness of 70-80 nm were prepared for electron microscopy and examined with an FEI Morgagni 268 Transmission Electron Microscope operating at 80kV. Electron micrographs were obtained using the Olympus-SIS Morada digital camera (Olympus Soft Imaging Solutions, Münster, Germany).

### RNA-sequencing (RNA-seq)

Total RNA was extracted from patient F3-II:1 and control skeletal muscle biopsies (∼15-50 mg) using the RNeasy Fibrous Tissue Mini Kit (Qiagen, Hilden, Germany) according to the manufacturer’s instructions. Strand-specific Poly-A+ RNA libraries were prepared from extracted RNA using the Agilent SureSelect XT library preparation kit (Agilent, Santa Clara, CA, USA). QC was performed using TapeStation 4200 (Agilent, CA, USA) and Qubit 4 Fluorometer (Thermo Fischer Scientific, Waltham, Massachusetts, USA), as well as QC sequencing on an Illumina iSeq 100 flowcell (Illumina, San Diego, CA, USA). These strand-specific libraries were sequenced on an Illumina NovaSeq 6000 to produce paired-end 150 bp reads and an average of 50 million read pairs per sample. Adaptor sequences were removed and demultiplexed FASTQ files were provided by Genomics WA (Western Australia) for download and further analysis. FASTQ files were processed, including read quality control and alignment, using the nf-core/rnaseq pipeline (https://nf-co.re/rnaseq), v3.8.1. Trimmed reads were aligned to the NCBI GRCh38 human reference genome using STAR v2.7.10a (13) (STAR, RRID: SCR_004463). We used DROP v1.0.3 (14), as previously described (15) to analyse aberrant gene expression amongst a cohort of 129 skeletal muscle RNA-seq datasets from rare muscle disease patients and unaffected controls. DROP leverages OUTRIDER (16), which uses a denoising autoencoder to control co-variation before fitting each gene over all samples via negative binomial distribution. Multiple testing correction was done across all genes per sample using DROP’s in-built Benjamini-Yekutieli’s false discovery rate (FDR) method. Plots were prepared using R (v4.1.3) in RStudio. The splicing pattern and expression of *JPH1* in F3 was visualised using Integrative genomics Viewer (IGV) (17) and plots were created using ggsashimi (18).

### Data Sharing statement

ES and srGS data of probands and family members is available on seqr. All relevant clinical data is shared as part of this study.

Identified variants in JPH1 have been submitted to ClinVar with accession numbers SCV004228294 - SCV004228296.

Code for generating plots is available at: https://github.com/RAVING-Informatics/jph1-cm

## Results

### Clinical findings in patients with *JPH1*-related myopathy

The clinical findings of all three probands are summarised in Table 1. In general, the three probands had a remarkably similar presentation with marked facial and bulbar weakness, in addition to respiratory involvement, limb muscle weakness and generalised muscle wasting. Facial weakness was accompanied by bilateral ptosis and ophthalmoplegia (patient F2 II.1, Fig 1b), a nasal voice and dysphagia. They also presented with myalgia, exercise intolerance and fatiguability. Reduced forced vital capacity (FVC) was prominent in F1-II:1 (19% of predicted value). The patients also showed lordosis and scoliosis (patient F2 II.1, Fig 1c). None of the patients showed any cardiac involvement or intellectual impairment.

**Table 1:** Clinical, histopathological, and MRI details of patients included in the study. Abbreviations used: NSAA=North Star Ambulatory Assessment, N/A = Not assessed, EM = Electron Microscopy, FVC = forced vital capacity, LL = lower limb, PEmmax and PImmax = maximal static inspiratory and expiratory pressures, NGT = nasogastric tube, UL = upper limb.

| Patient ID | F1-II:1 | F2-II:1 | F3-II:1 |
| --- | --- | --- | --- |
| **JPH1 variant** | c.373del, p.(Asp125Thrfs*30) - homozygous | c.354C>A, p.(Tyr118*) - homozygous | c.1738del, p.(Leu580Trpfs*16) - homozygous |
| **Current age/age at last exam** | Fourth decade/Fourth decade | Deceased during late fifth decade /late fifth decade | Second decade/second decade |
| **Age of onset** | Congenital | Congenital | Congenital |
| **Motor development** | Not sitting during first year | Delayed motor milestones - walked at 0-5 yrs. | Delayed motor milestones. |
| **Progression** | Slowly progressive | Stable through childhood, slow decline since fourth/fifth decade. | Relatively stable through childhood |
| **Maximum motor ability** | Walking independently for up to 10 minutes | Able to walk unaided, able to-climb stairs with aid of railing. | Current motor ability |
| **Motor ability** | Mainly manual wheelchair user, able to stand up with assistance, unable to walk. | Able to stand up with assistance, able to walk 10 m with support. | Can walk around school, can run - but slow. Climbs stairs with railings. NSAA = 31/34 |
| **Muscle strength (MRI scale)** | No recent assessment. | Scapular 3/5, hip girdle 3/5, and distal UL and LL 4-/3. Unable to lift upper limbs to head, cannot tip-toe or stand on heels. | Hip girdle 4-/5, distal UL 4+/5, 4/5 power elsewhere, 1 - 2 handed Gowers. |
| **Distribution of weakness** | Generalised (proximal and distal, upper, and lower limbs), including facial weakness | Generalised (proximal and distal upper and lower limbs), including ptosis and facial and neck flexor weakness. | Generalised with ptosis and facial diplegia, no neck flexor weakness |
| **Reflexes** | Absent | Absent | Absent in LL; reduced in UL |
| **Contractures** | Talipes at birth. | Talipes at birth; ankle contractures. | No - hypermobile at ankles. |
| **Spine involvement** | Kyphoscoliosis, rigid spine. | Dorsal scoliosis, lumbar lordosis, and winged scapulae. | Mild lumbar lordosis; no scoliosis. |
| **Muscle wasting** | Generalised wasting | Generalised wasting | Generalised wasting |
| **Ocular symptoms** | Bilateral ptosis and ophthalmoplegia. | Ptosis and ophthalmoparesis (surgical correction of ptosis and squint in childhood). | Ptosis and ophthalmoplegia with mild limitation of upward gaze. |
| **Bulbar symptoms** | Dysphagia | Nasal voice, no dysphagia. | Mild dysarthria, Short term NGT feeds at birth. |
| **Respiratory involvement** | Respiratory insufficiency at birth<br>FVC=0.5L (19%). | No respiratory support. FVC=1.76L (63%).<br>Plmmax: 43%, PEmmax: 72%. | No respiratory support. FVC=1.22L (57%),<br>FEV1=1.20 (62%). Normal sleep study. |
| **Other** | Cleft palate, migraine. | Cleft palate. Myalgia and exercise intolerance. Hypoacusia in the fourth decade (neural deafness). Alcoholic liver cirrhosis, died from hepatic insufficiency (transplant excluded). | Exercise intolerance with fatigue. |
| **EMG findings** | Mild myopathic changes (in childhood) | Myopathic features. Normal RNS and jitter. | N/A |
| **Muscle histopathology** | Biopsy in second decade, quadriceps. Type 1 myofibre predominance, <2% of myofibers are type 2. Some hypertrophied myofibres and disseminated atrophic myofibres of both types. | Biopsy in second decade, deltoid. Type 1 myofibres predominance, few centralised nuclei. Occasional hypertrophic myofibres and scattered hypotrophic type II fibers. Mild focal fibrosis. No myofibre degeneration/regeneration, nor abnormalities on oxidative reactions. | Biopsy of upper arm. Type 1 myofibre predominance and most of the atrophic myofibres are type 2, there is no grouping of myofiber types. |
| **EM findings** | N/A | N/A | Slightly reduced triads, occasional small minicore-like foci. |

### Identification of deleterious variants in *JPH1*

Analysis of ES (F1 and F2) and srGS (F3) data were initially negative for approximately 600 genes known to cause a neuromuscular phenotype (19, 20). Subsequently, we identified three unique homozygous protein truncating variants in *JPH1*: two in exon 1 (c.373del, p.(Asp125Thr*fs*\*30) and c.354C>A, p.(Tyr118*), in F1 and F2 respectively and one in exon 4 (c.1738del, p.(Leu580Trp*fs\**16) in F3 (Fig. 1d-e). Using VarSome and Alamut we assessed the pathogenicity of the identified variants. Since all three variants would result in null alleles and were absent in the reference population databases, they fulfilled the PVS1 (very strong), PM2 (supporting) and PS3 (strong) criteria of ACMG guidelines, resulting in classification of the variants as “Likely pathogenic/Pathogenic”. Sanger sequencing of the available family members supported the co-segregation of the bi-allelic *JPH1* variants with the clinical presentation in the three families (Fig. 1d).

### Muscle pathology associated with biallelic loss-of-function *JPH1* variants

Muscle biopsies from patients F2-II:1 and F3-II.1 revealed a striking pattern of type 1 myofiber predominance (Fig. 2b). No other major features were observed, there was no increase in internal or centrally located nuclei, there was no staining suggestive of cores or nemaline bodies. Electron microscopy analysis of the muscle biopsy of F3-II:1 showed ultrastructural defects including rare examples of Z-band streaming, slightly reduced number of triads and structurally abnormal sarcoplasmic reticulum which appeared dilatated (Fig. 2d-f).

**Fig. 2.**
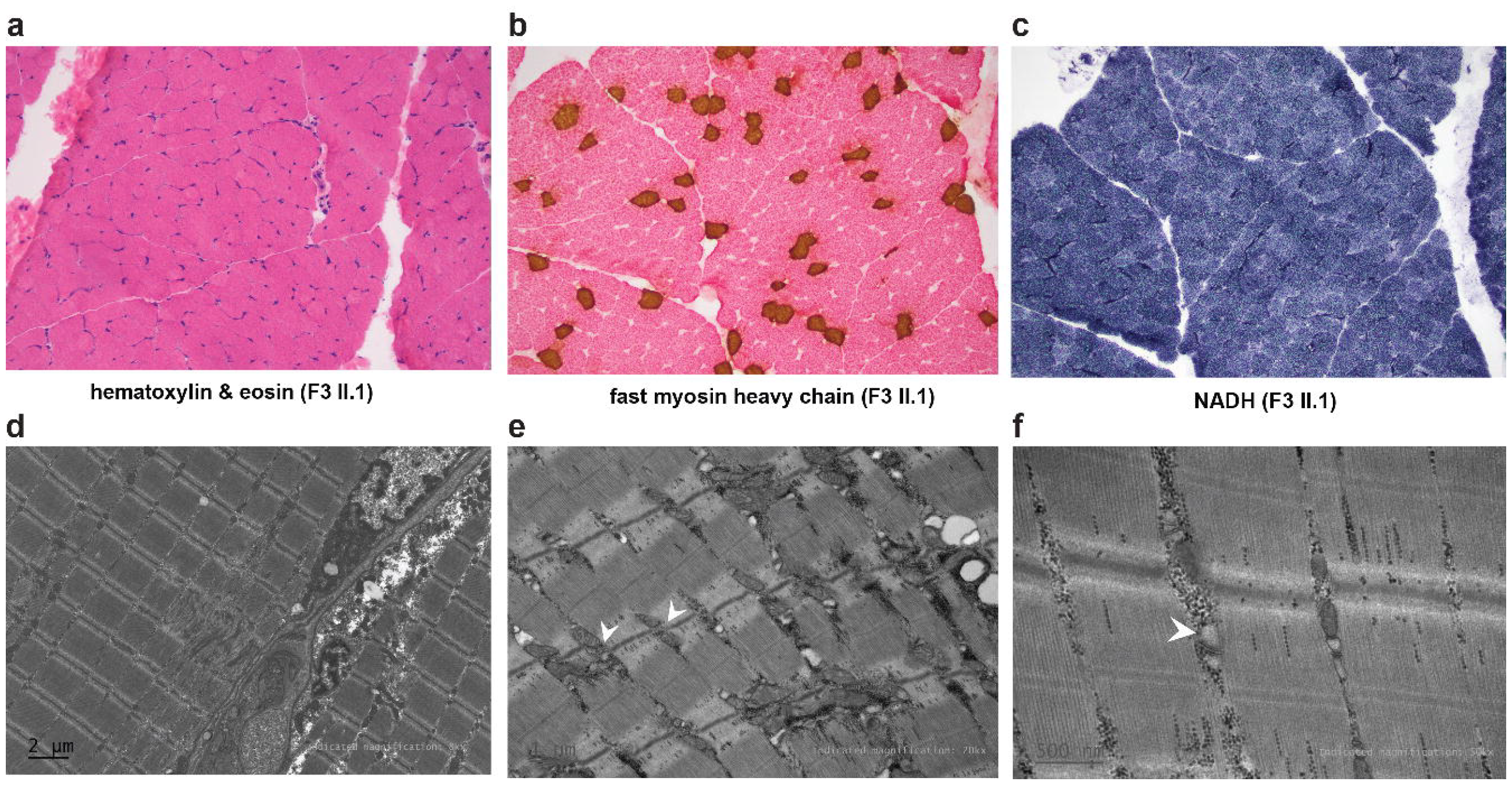
Muscle pathology of patient F3 II.1. a) Hematoxylin & eosin (HE) showing preserved muscle structure. b) IHC for fast myosin heavy chain (MHC; stained with DAB (brown)) and eosin showing type 1 myofibre predominance. Most of the atrophic myofibres stain as type 2 but the normal chequerboard distribution of fibre types appears relatively preserved c) NADH staining. No cores, or mini cores are present. Electron microscopy (patient F3 II.1) showing evidence of focal Z-band streaming d), and reduced triads with dilatated SR (white arrowheads) e-f).

### Analysis of skeletal muscle RNA-seq data from proband F3-II:1

OUTRIDER analysis detected under expression of *JPH1* as an outlier (Fig. 3a) in F3-II:1 (Z=-8.48, p-adj=1.15 x 10^-8^). Based on normalised gene counts, *JPH1* expression in F3-II:1 was the lowest at 1167.87 compared to the other 129 muscle disease patients and healthy controls (log_2_fold change = -2.84). IGV analysis (Fig. 3a) and sashimi plots (Fig. 3c) confirmed the low expression of gene.

**Fig. 3.**
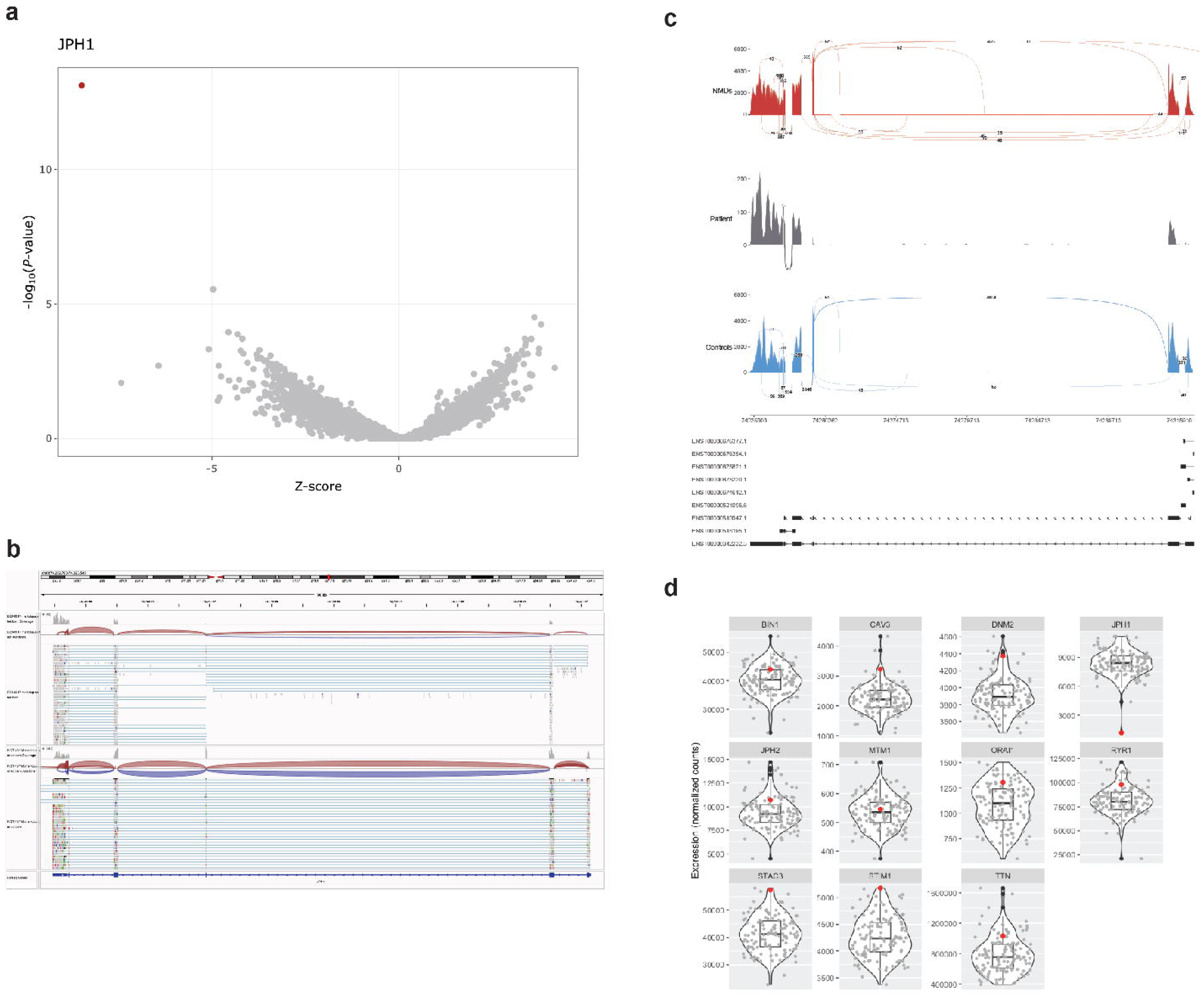
Splicing and gene-expression in skeletal muscle from a patient with *JPH1*-related myopathy. a) Volcano plot showing results from the OUTRIDER analysis. *JPH1* is indicated as an outlier in red colour. b) Visualisation of RNAseq data in IGV comparing *JPH1* expression with a control (we need to change the names of the patient and control bam files in the final image). c) *ggsashimi* plot analysis of *JPH1* from RNAseq data of F3-II:1. *JPH1* expression and splicing patterns of the patient are shown in grey colour, compared with other NMD patients in red (n=39) and unaffected controls in blue (n=6). d) Normalised expression of genes encoding for other triad proteins is presented as boxplots. Median and quartile values are shown, with whiskers reaching up to 1.5 times the interquartile range. Expression levels from individual samples in the cohort are shown with jitter points and that of F3-II:1 is represented with red colour. The violin plot illustrates the distribution of data in each cohort. The scaled Y-axis shows normalised counts.

Since, a reduced expression of *JPH1* could affect other T-tubule proteins, we additionally analysed the expression of other genes encoding components of the triad and T-tubules, including: *RYR1, CACNA1S, JPH2, MTM1, DNM2, BIN1, STAC3, ORAI1, STIM1, CAV3* and *TTN* (Fig. 3d). There were no differences in expression levels for any of these genes of interest in *JPH1*-related myopathy compared to healthy control muscle or patients with other forms of neuromuscular diseases.

## Discussion

Our results demonstrate that loss of function variants in *JPH1,* coding for Junctophillin-1, result in a novel congenital myopathy, characterised by global distribution of muscle weakness and wasting, but with prominent facial muscle weakness, bilateral ptosis, exercise intolerance and fatiguability.

In skeletal muscles, junctophilins have a regulatory and maintenance function with other triad proteins, including assembly of Ca^2+^ release complex and organisation of the Store Operated Ca^2+^ entry (SOCE) pathway through interaction with other T-tubule proteins including RYR1, DHPR and CAV3 (2).

All three probands showed prominent myalgia, along with exercise intolerance and fatiguability. These features are commonly seen in other triadopathies, such as tubular aggregate myopathies caused by pathogenic variants in *STIM1* (3).

In our patients we observed homozygous null variants in *JPH1* resulting in no expression of complete transcript indicating no viable production of JPH1. This is well reflected in our morphological and ultrastructural studies which concur with *Jph1* KO mice. EM analysis of muscle biopsy of F3-II:1 showed reduced number of triads. Light microscopy analysis of F2-II:1 and F3-II:1 showed predominance of type 1 myofibres. Generally, triad abundance varies in different myofibre types in skeletal muscle. Type 1 myofibre predominance is also observed in other triadopathies, including *RYR1, DNM2, BIN1*, and *MTM1*-associated congenital myopathies (21-24).

EM analysis of muscle biopsy of F3-II:1 also showed dilated SR. Disorganisation of triads and swelling of SR was observed in mutant muscles of *Jph1* KO mice (4). The swelling of SR can be attributed to SR Ca^2+^ overloading and has been seen in mice lacking both *ryr1* and *ryr3* (25). Hence, in human muscles lacking JPH1, like mice, SR Ca^2+^ overloading could cause similar abnormalities due to reduced triad junctions potentially hindering DHPR-mediated activation of RYR.

Congenital myopathies arising due to pathogenic variants in genes encoding components of the triads or proteins involved in triad formation and maintenance, including RYR1 and STAC3, share many clinical features. (26, 27).. These include hypotonia and axial weakness, which often tends to be static or slowly progressive, facial and bulbar weakness, resulting in dysphagia and dysarthria, ocular weakness, including ptosis and ophthalmoplegia and respiratory insufficiency. Joint contractures may be present at birth (26, 27).

Previously, deficiency of Jph1 in mouse models was shown to result in neonatal death. This was attributed to failure in suckling, as a newborn due to weak contractile activity of jaw muscles and weak pharyngoesophageal or diaphragm muscles (4). The myofibers of these *Jph1* KO mice were morphologically normal, and analysis of muscle histology did not detect obvious abnormalities. Ultrastructural analysis using electron microscopy, however, revealed that *Jph1* KO neonates had swollen SR and defective and highly reduced triads. These observations suggested that loss of JPH1 clearly affects triad formation in skeletal muscles (4).

Additionally, reduced JPH1 expression has been associated with defective triad formation and disturbed Ca^2+^ homeostasis due to mislocalisation of RYR1 and DHPR (7, 8, 28). While the disease presentation is similar in *Jph1* KO mice and *JPH1* patients, none of our patients had severe muscle weakness or a dystrophic phenotype as seen in neonatal mice.

Analysis of RNA-seq data showed that mRNA expression of other key genes of the triad are unaltered in *JPH1* patient skeletal muscle biopsy compared to healthy controls and other neuromuscular disease biopsies. This is perhaps not surprising given the relatively mild phenotype observed in these patients, compared to affected individuals with bi-allelic loss-of-function variants in *CACNA1S, RYR1* or *STAC3*.

This would suggest that the loss of JPH1 observed in our patients due to homozygous null variants, affect the triad formation and maintenance. The exact pathomechanism of how loss of JPH1 and normal expression of other triad genes contribute to the phenotype, remains to be understood.

Our results, for the first time show that bi-allelic null variants in *JPH1* cause a congenital myopathy with facial and ocular muscle weakness. Hence, *JPH1* should be included in genetic screenings of unsolved patients with similar clinical presentation.

## Data Availability

Data Sharing statement
ES and srGS data of probands and family members is available on seqr. All relevant clinical data is shared as part of this study.
Identified variants in *JPH1* have been submitted to ClinVar with accession numbers SCV004228294 - SCV004228296.
Code for generating plots is available at: https://github.com/RAVING-Informatics/jph1-cm

## Author contributions

Conceptualisation of the study: GR, VS

Project administration: GR, VS

Funding acquisition: MJ, AT, GR, VS

Supervision: GR, VS

Patient samples and data collection: AC, TR, JJV, PM, EH, CMB

Data analysis and curation: MJ, AT, CF

Methodology: MJ, AT, CF, LD, JD

Visualisation: MJ, CF

Writing the original draft: MJ, AT, CF

Review and editing of the manuscript: MJ, AT, TR, AC, EH, JJV, GR, VS

## Acknowledgements

Sequencing was conducted in the Genomics WA Laboratory in Perth, Australia. BioPlatforms Australia, State Government Western Australia, Australian Cancer Research Foundation, Cancer Research Trust, Harry Perkins Institute of Medical Research, Telethon Kids Institute, and the University of Western Australia support this facility. We gratefully acknowledge the Australian Cancer Research Foundation and the Centre for Advanced Cancer Genomics for making available Illumina Sequencers for the use of Genomics WA.

